# Identifying patients with a phenotype consistent with chronic postsurgical pain after hip and knee arthroplasty using robust, scalable k-medoids clustering analysis

**DOI:** 10.64898/2026.08.11.26360161

**Authors:** Lawrence Gillam, Brett Doleman, Roger Knaggs, John Williams

## Abstract

**Background:** Chronic postsurgical pain (CPSP) affects between 7-23% and 13-44% of patients after hip and knee arthroplasty, respectively. Standardised methods of pain assessment provide superior evaluation of pain, including the Oxford Joint Score Pain Subscale (OJS-PS). We aim to estimate the proportion of patients with a phenotype consistent with CPSP through a k-medoids clustering technique and identify a threshold on the OJS-PS to highlight such patients at a population level.

**Methods:** In this cross-sectional study Patient Reported Outcomes Measures data 6-months after hip and knee arthroplasty from 2017 to 2025 were examined. An adapted *k*-medoid clustering technique utilising subsampling, batch assignment and probabilistic consensus allocated clusters. A receiver operator characteristic analysis identified a threshold on the OJS-PS noting the lowest scoring cluster. Our categorisation was compared to self-reported severe or moderate pain; sensitivity, specificity and accuracy of this categorisation were calculated.

**Results:** We analysed 109,542 hip and 113,799 knee arthroplasty patients; three clusters were used in each analysis. After hip arthroplasty: 14.4% of patients were assigned to the cluster with the lowest median OJS-PS of 11 [IQR 8 – 13]. A threshold of 15.5 classified patients as severe or moderate pain with 60.6% sensitivity, 91.0% specificity and 85.7% accuracy. Similarly, after knee arthroplasty, 25.3% were assigned to the cluster with the lowest median OJS-PS of 14 [IQR 11 – 16]. A threshold of 18.5 on the OJS-PS had an 85.4% sensitivity, 88.4% specificity and 87.8% accuracy for classifying patients with self-reported severe or moderate pain.

Conclusions

This robust and scalable clustering technique on ordinal clinical data estimates the proportion of patients reporting a phenotype consistent with CPSP. On a population level the thresholds identified on the OJS-PS could aid screening for potential CPSP patients 6 months after hip and knee arthroplasties.

## Introduction

Hip and knee arthroplasties are among the most common operations performed annually (1, 2). They are primarily performed in patients with end-stage osteoarthritis to improve pain, function and quality of life (3, 4) where other management options have not been successful.

Potential complications following either hip or knee arthroplasty, such as death, deep vein thrombosis, myocardial infarction or infection occur in 1 – 3% of cases (5–8). In contrast, chronic postsurgical pain (CPSP), i.e. *‘pain that develops or increases in intensity after a surgical procedure or a tissue injury and persists beyond the healing process, i.e. at least 3 months after the surgery’* (9) has an estimated prevalence of between 7 to 23% after hip and 13 to 44% following knee arthroplasty (10). This can be impactful on an individual’s life, with deleterious effects on quality of life, mood, function, analgesic requirement as well as on the wider health care system (10–14). Yet, patients suffering with CPSP may not present to health services (15), therefore their illness may go unrecognised and untreated.

Asking a patient to rate their pain is subjective and one-dimensional and may exhibit recency bias or follow the peak-end rule (16). Therefore, in chronic pain research, a variety of scales have been developed (17). The Oxford Joint Score (OJS) is a scoring system used to assess patients following a hip and knee arthroplasty, this can be divided into pain subscale (OJS-PS) and functional domains (18). This standardised survey provides a more comprehensive assessment of a patient’s pain and is less prone to bias. Being able to timely identify a patient suffering with CPSP based on these questions has high utility and could improve patients’ outcomes. However, this relies on a standardised and repeatable way to identify such patients and having a threshold in which chronic pain patients are identified.

There were three aims of this work:

1. Report the proportion of patients with a phenotype consistent with CPSP at 6 months in England following an elective hip or knee arthroplasty
2. To apply a robust and scalable k-medoids clustering technique on a large publicly available dataset to identify different patient clusters.
3. Identify a threshold on the OJS-PS that can be used to screen for patients with a phenotype consistent with CPSP after hip and knee arthroplasty.

## Methods

### Dataset

The Patient Report Outcomes Measures (PROM) published by NHS England from the financial year April 2017 to March 2025 were analysed. This is an anonymised patient level dataset collected prior to and at 6 months after the surgery providing information on age, co-morbidity, pain and functional status both pre- and postoperatively. Revisional surgeries were excluded.

These datasets and method of collection are publicly available via the NHS Digital platform (19). The R-script and code for the assimilation of these datasets is provided in the supplementary information, as is the dataset used for this analysis.

### Oxford Joint Score: Pain Subscale

The OJS is a 12-item survey. Each question in the OJS is scored on a Likert scale from 0 to 4, with a score of zero indicating greatest severity, and a score of 4 indicating no problem. However, this survey includes questions not directly associated with pain. *Harris et el.,* proposed a subscale of this measure that can better assess an individual’s pain (18, 20).

The Oxford Knee Score Pain Subscale (OKS-PS) has 7 questions (20), whilst the Oxford Hip Score Pain Subscale (OHS-PS) has 6 questions (18). The pain subscale scores were calculated from the PROM dataset. All questions and answers are provided in the supplementary information. This pain subscale has been used in previously published work (21).

### k-Medoid Cluster Analysis

Results of the OJS-PS are provided on a 5-point Likert scale with answers such as ‘mild’, ‘moderate’ and ‘severe’ provided. These are then numerically coded. These are ordinal rather than continuous interval in nature. Conventional k-means clustering uses Euclidean distance, therefore is most appropriate for continuous interval or ratio data. These assumptions cannot be guaranteed when applied to questions composing the OJS-PS. Consequently, the ‘*Gower’* distance was used with k-medoids clustering as a preferred clustering technique, and later with confirmatory hierarchical clustering.

For clarity, cluster assignment is based on the pattern of response to the 6 or 7 questions within the OJS-PS, rather than the overall summed OJS-PS score. This maintains the multidimensionality of the pain assessment.

The dataset was scaled and only complete observation were analysed, imputation was used if more than 5% of data was missing (22, 23). Initially, a clustered heatmap was produced to demonstrate the postoperative reporting of questions in the OKS-PS and OHS-PS.

The number of clusters was determined, *k*, through assessment and balancing of the clustering statistics, including gap and silhouette statistic as well as though using the elbow method (*Supplementary Information*). There was no pre-specified single statistic that dictated the cluster number (24, 25). The data was divided into *k* clusters. Each patient was assigned to a cluster. Therefore, the OKS-PS and OHS-PS of each cluster was explored.

An adapted method proposed by *de Mathelin et al* (26) was utilised:

1. Initial k-medoid assignment on a sample of 5% of patients assigned to precalculated number of clusters (*k*).
2. Datapoints which were not within this initial sample were then batch-assigned to a cluster which they most closely represented in accordance with the partitioning around medoid (PAM) clustering algorithm. This was repeated until all data points had an assigned cluster. This approach has been shown to give reliable results when compared to other clustering methods whilst limiting the computation cost (26).
3. As the clustering algorithm is sensitive to initial conditions and random sampling, this process was repeated up to 100 times and cluster assignment aligned per iteration.
4. After each iteration the probability of being assigned to an individual cluster was updated. A stopping condition was made so that only if the difference in probability of cluster assignment was less than 0.001 on 10 consecutive iterations would the algorithm stop.

The descriptive statistics of the OJS-PS per cluster are provided. The cluster that had the lowest average OJS-PS represented a cluster of patients with the most severe pain and/or functional limitation. Therefore, referred to as the ‘*chronic pain cluster’: being assigned to this cluster does not constitute a diagnosis of CPSP, rather such patients are likely to have a phenotype consistent with CPSP*. We report the demographic details by each cluster, including age, sex, pre-operative living arrangement as well as noting patients reporting of postoperative pain, average responses of questions making up the OJS-PS, postoperative EQ5D quality of life measure, median postoperative OJS-PS, full OJS and mean change in OJS (postoperative score minus preoperative score). Finally, we report the patient self-reported level of success and satisfaction of the operation.

A threshold on the OJS-PS was determined using a Receiver Operator Characteristic (ROC) Curve to highlight the cluster with the lowest OJS-PS scores, as this is indicative of worse pain and/or functional limitation.

### Internal Consistency

To assess the ability of our threshold to categorise chronic pain patients we examined the number and proportion of patients that report ‘postoperative pain’ at 6-months. A two-by-two table was created based on the OJS-PS being above or below our threshold, and whether a patient reported moderate or severe pain compared to mild, very mild or no pain. We calculated the sensitivity, specificity and accuracy with 95% confidence interval for this categorisation. Finally, a summarising heatmap of the components of the OJS-PS is provided but categorised according to the threshold identified in this analysis to give readers an appreciation of the overall reporting that patients give when their postoperative OJS-PS is lower than our threshold. Hierarchical clustering was used as an internal validation tool to assess results obtained from the k-medoids clustering.

### Ethical Statement

As this work was a secondary analysis of anonymised, publicly available data, with no direct patient involvement formal ethical approval was not required.

### Statistics and Code

Analyses were conducted using R-Studio (2026.01.0+392); all code is available in supplementary information. Descriptive data are presented as the median with [interquartile range] provided unless stated otherwise, sensitivity, specificity and accuracy are presented as a percentage with 95% confidence intervals. A Chi-Squared test was used to assess differences across categorical variables, and an ANOVA was used to compare continuous data between groups. Statistical significance was set at P<0.05.

STROBE guidelines were adhered to in this work.

## Results

The patient reported outcome measures data published by NHS Digital from financial year April 2017 to March 2025 contains data from 109,542 patients from elective, non-revisional, hip arthroplasties and 113,799 patients following elective, non-revisional, knee arthroplasties. Within the ‘hip’ dataset there were 1,272 entries with missing data (1.16%) related to the OHS-PS. In the knee dataset there were 373 missing entries (0.33%) associated with the OKS-PS. Our plan to utilise imputation techniques was precluded due to the low proportion (<5%) of missing data. The patients with missing data were excluded from our analysis.

Baseline characteristics were comparable between both hip and knee cohorts (Table-1). Note that within the knee arthroplasty dataset there was no recording of stroke history, nor were any patients aged between 20 and 40.

**Table-1.**
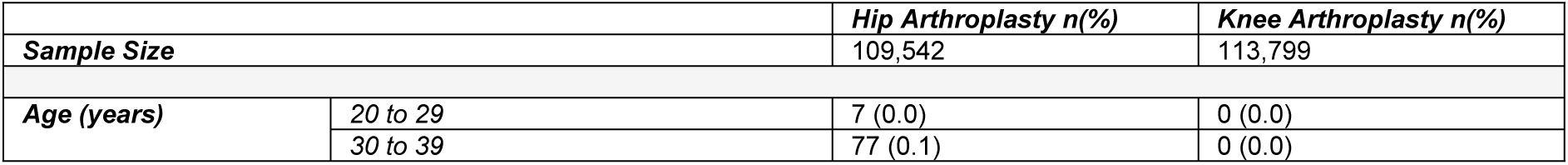
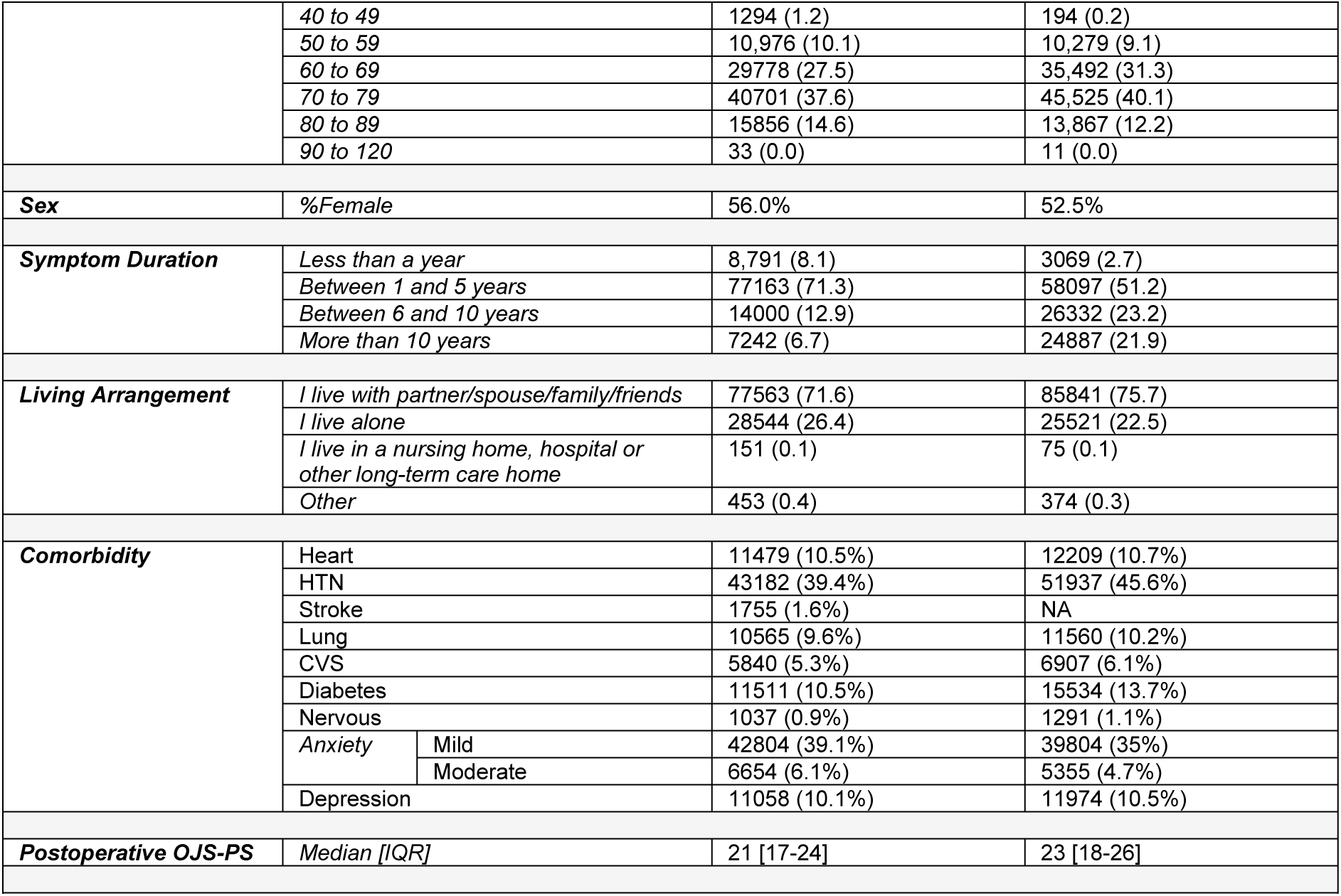
Summary of demographic data of non-revisional hip and knee arthroplasties from 2017 to 2025.

Postoperative OJS-PS scores for both hip and knee arthroplasties show a negative skew. Across the entire dataset the median OHS-PS was 21 [17 – 24] and OKS-PS 23 [18 – 26]. Self-reported pain reporting general correlated with OJS-PS following both hip and knee arthroplasty (Table-2).

**Table-2.** A summary of the Oxford Joint Score – Pain Subscale following both hip and knee arthroplasty categorised by patient self-reporting of pain severity.

|  | Oxford Hip Score – Pain Subscale (Maximum = 24) |  |  |  |  | Oxford Knee Score – Pain Subscale (Maximum = 28) |  |  |  |  |
| --- | --- | --- | --- | --- | --- | --- | --- | --- | --- | --- |
| Pain Severity Reported | Median | Lower Quartile | Upper Quartile | Min | Max | Median | Lower Quartile | Upper Quartile | Min | Max |
| Severe | 16 | 9 | 19 | 0 | 20 | 7 | 4 | 10 | 0 | 24 |
| Moderate | 13 | 9 | 16 | 1 | 21 | 15 | 11 | 17 | 1 | 25 |
| Mild | 16 | 14 | 18 | 3 | 22 | 20 | 17 | 22 | 2 | 26 |
| Very Mild | 20 | 18 | 22 | 3 | 23 | 24 | 22 | 26 | 3 | 27 |
| None | 24 | 23 | 24 | 4 | 24 | 28 | 26 | 28 | 5 | 28 |

### Hip and Knee Analysis

A heatmap showing answers to the OHS-PS and OKS-PS was used to visualise data (Fig 1 and 2).

**Fig-1.**
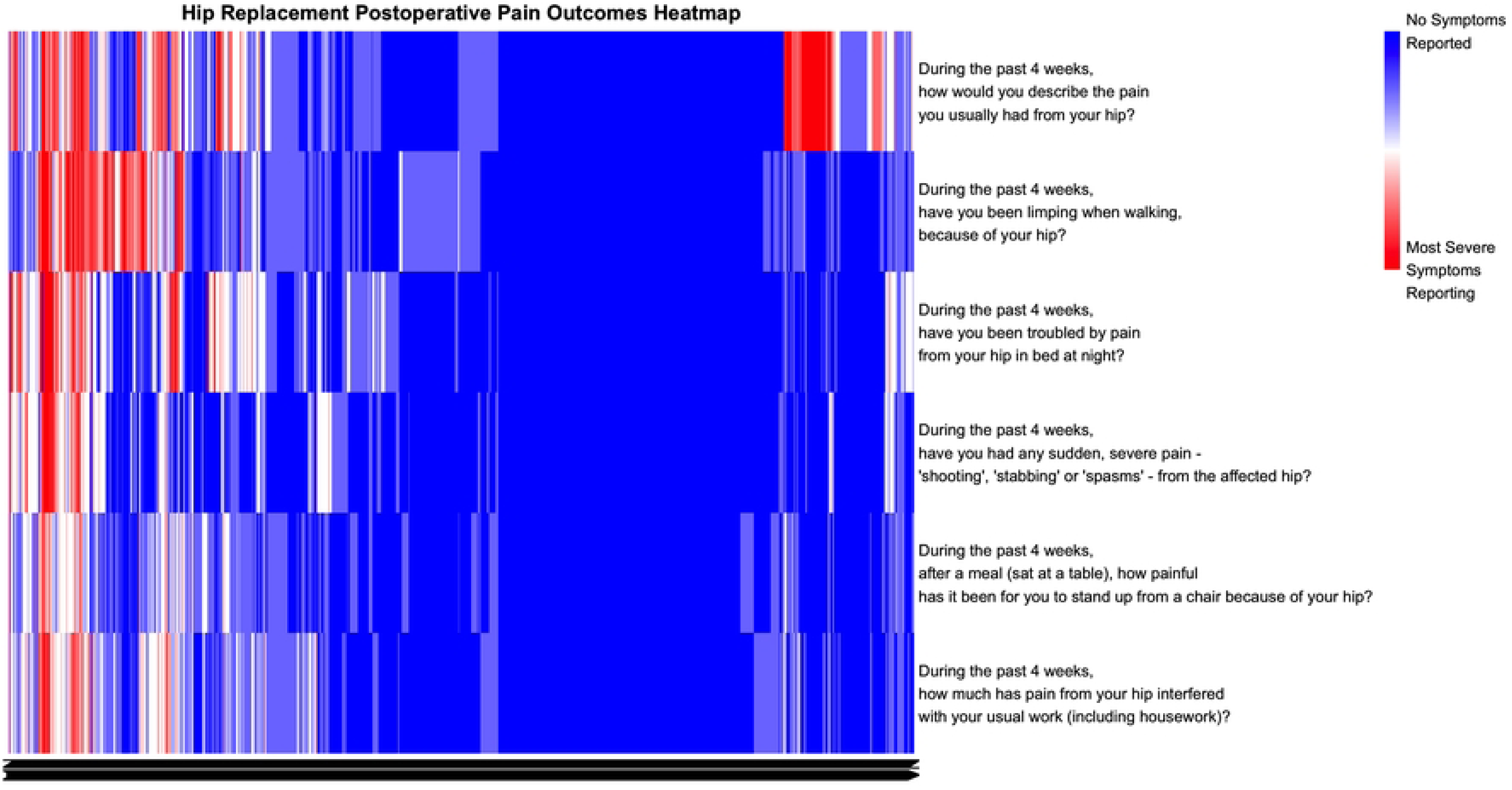
Heatmap of answers to the Postoperative Oxford Hip Score – Pain Subscale. Each question asked as part of the score is represented by each row. The answers provided are on a scale from 0 to 4, with 0 representing the most severe symptoms/issues, and 4 representing no symptoms/issues, this is shown via a change in colour from red to blue.

**Fig-2.**
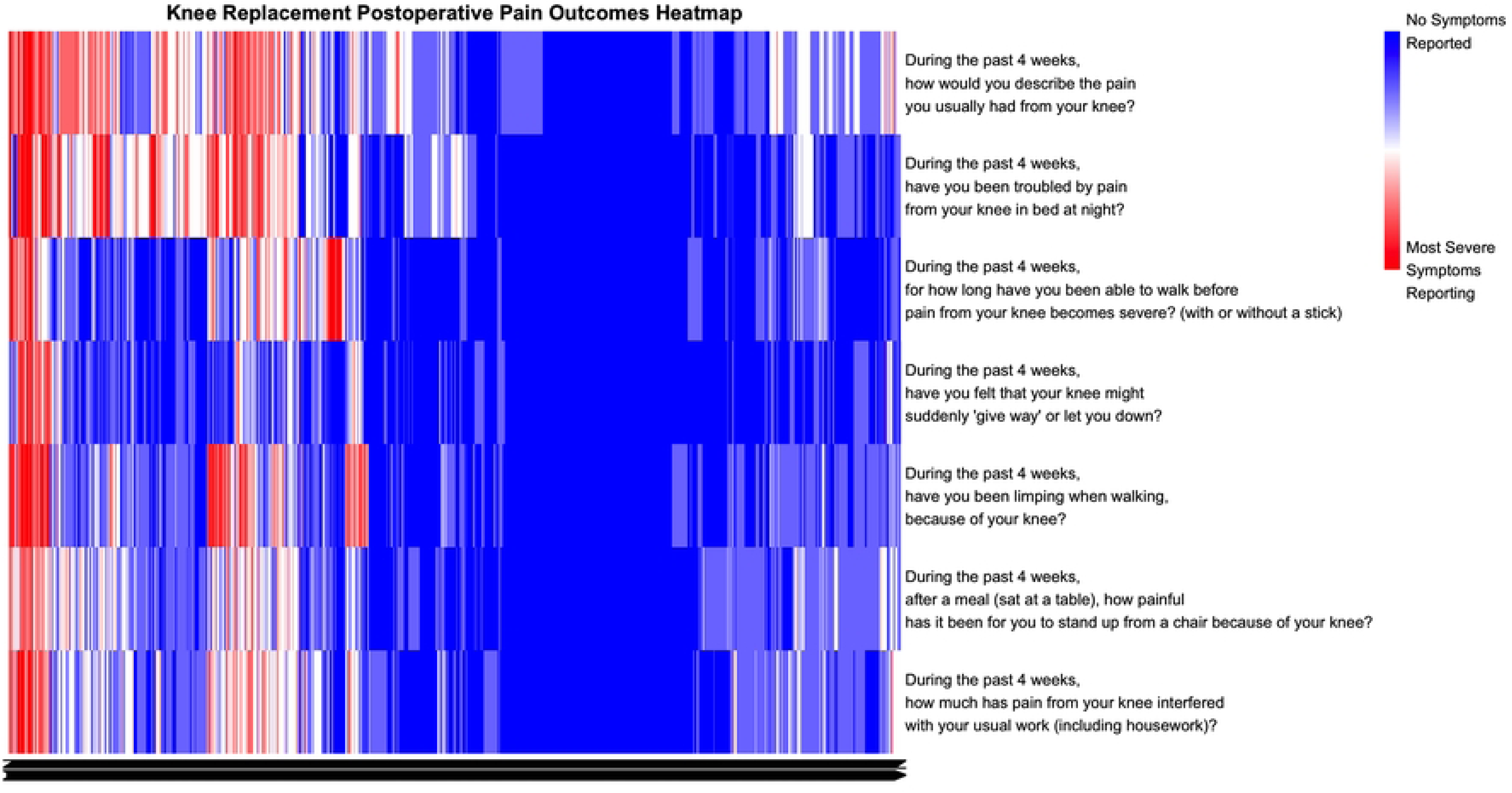
Heatmap of answers to the Postoperative Oxford Knee Score – Pain Subscale. Each question asked as part of the score is represented by each row. The answers provided are on a scale from 0 to 4, with 0 representing most severe symptoms/issues, and 4 representing no symptoms/issues, this is shown via a change in colour from red to blue.

### k-Medoid Clustering

There was no unique optimum cluster number identified through cluster statistics. Cluster number was decided on through using the gap and silhouette statistics with the elbow method. It was concluded at a *k*=3 for both the hip and knee cohort would balance the statistical outputs with clinical interpretability (*Supplementary Information: S1 Fig-1, S1 Fig-3, S1 Table-1 and 2*). Following division into three clusters, the median OHS-PS for each cluster was: *cluster-1:* 23 [22 – 24], *cluster-2:* 18 [17 – 20] and *cluster-3* of 11 [8 – 13]. We found that 14.4% of the hip cohort were assigned to *cluster-3*, which we refer to as the ‘*chronic pain cluster*’ (Table-3). After division of the knee cohort, the median OKS-PS of *cluster-1* was 21 [20 - 24], *cluster-2* was 27 [26 – 28] and *cluster-3* was 14 [11 - 16]. Within the knee cohort ‘chronic pain cluster’ there were 25.3% of patients (Table-3).

**Table-3.**
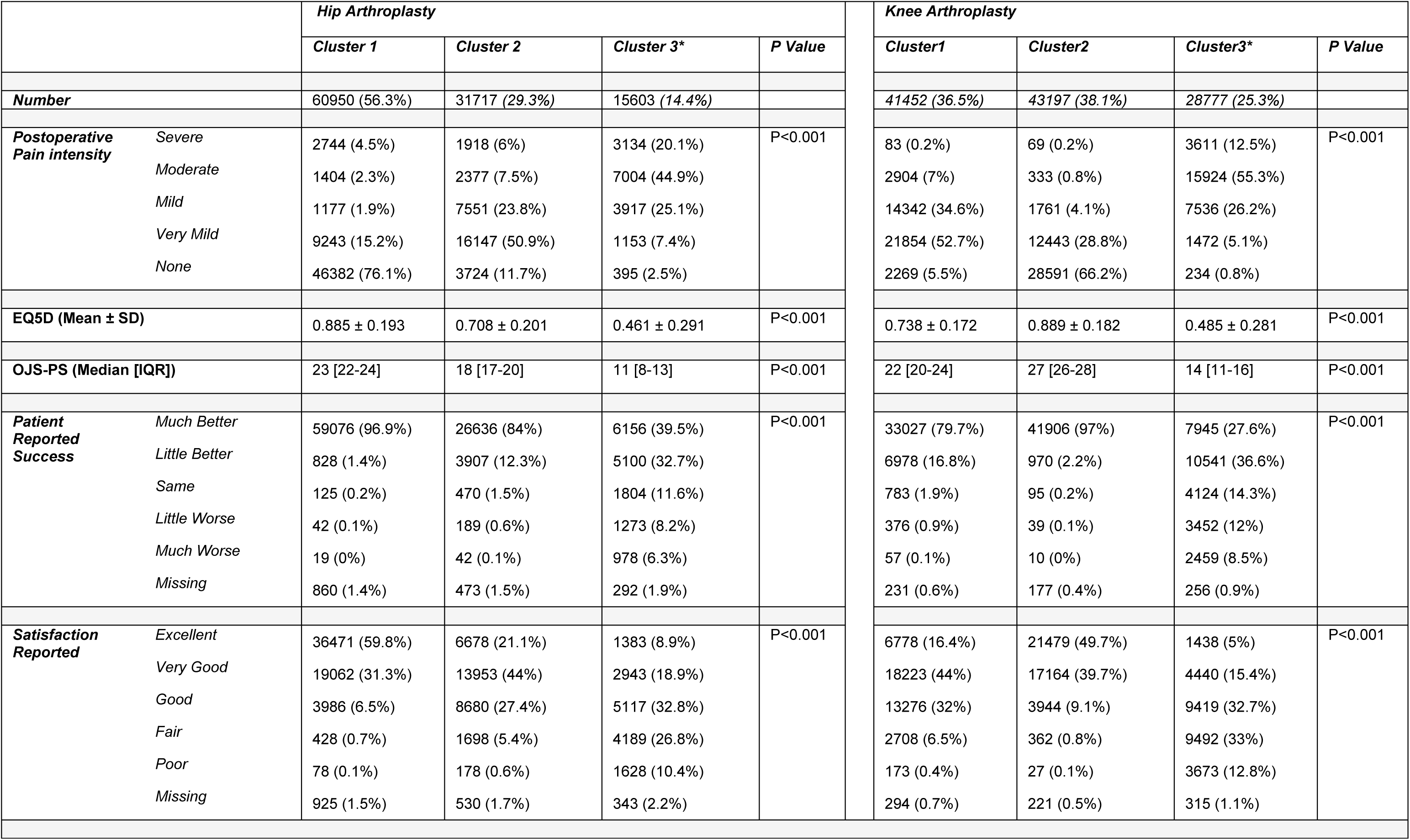
Summary of Pain reporting, Oxford Hip and Knee Score Pain Subscale, Quality of Life Measure (EQ5D), success and satisfaction by clusters formed through k-medoids clustering following arthroplasty. * = ‘chronic pain cluster’.

|  |  | Hip Arthroplasty |  |  |  | Knee Arthroplasty |  |  |  |
| --- | --- | --- | --- | --- | --- | --- | --- | --- | --- |
|  |  | Cluster 1 | Cluster 2 | Cluster 3* | P Value | Cluster1 | Cluster2 | Cluster3* | P Value |
| Number |  | 60950 (56.3%) | 31717 (29.3%) | 15603 (14.4%) |  | 41452 (36.5%) | 43197 (38.1%) | 28777 (25.3%) |  |
| Postoperative Pain intensity | Severe | 2744 (4.5%) | 1918 (6%) | 3134 (20.1%) | P<0.001 | 83 (0.2%) | 69 (0.2%) | 3611 (12.5%) | P<0.001 |
|  | Moderate | 1404 (2.3%) | 2377 (7.5%) | 7004 (44.9%) |  | 2904 (7%) | 333 (0.8%) | 15924 (55.3%) |  |
|  | Mild | 1177 (1.9%) | 7551 (23.8%) | 3917 (25.1%) |  | 14342 (34.6%) | 1761 (4.1%) | 7536 (26.2%) |  |
|  | Very Mild | 9243 (15.2%) | 16147 (50.9%) | 1153 (7.4%) |  | 21854 (52.7%) | 12443 (28.8%) | 1472 (5.1%) |  |
|  | None | 46382 (76.1%) | 3724 (11.7%) | 395 (2.5%) |  | 2269 (5.5%) | 28591 (66.2%) | 234 (0.8%) |  |
| EQ5D (Mean ± SD) |  | 0.885 ± 0.193 | 0.708 ± 0.201 | 0.461 ± 0.291 | P<0.001 | 0.738 ± 0.172 | 0.889 ± 0.182 | 0.485 ± 0.281 | P<0.001 |
| OJS-PS (Median [IQR]) |  | 23 [22-24] | 18 [17-20] | 11 [8-13] | P<0.001 | 22 [20-24] | 27 [26-28] | 14 [11-16] | P<0.001 |
| Patient Reported Success | Much Better | 59076 (96.9%) | 26636 (84%) | 6156 (39.5%) | P<0.001 | 33027 (79.7%) | 41906 (97%) | 7945 (27.6%) | P<0.001 |
|  | Little Better | 828 (1.4%) | 3907 (12.3%) | 5100 (32.7%) |  | 6978 (16.8%) | 970 (2.2%) | 10541 (36.6%) |  |
|  | Same | 125 (0.2%) | 470 (1.5%) | 1804 (11.6%) |  | 783 (1.9%) | 95 (0.2%) | 4124 (14.3%) |  |
|  | Little Worse | 42 (0.1%) | 189 (0.6%) | 1273 (8.2%) |  | 376 (0.9%) | 39 (0.1%) | 3452 (12%) |  |
|  | Much Worse | 19 (0%) | 42 (0.1%) | 978 (6.3%) |  | 57 (0.1%) | 10 (0%) | 2459 (8.5%) |  |
|  | Missing | 860 (1.4%) | 473 (1.5%) | 292 (1.9%) |  | 231 (0.6%) | 177 (0.4%) | 256 (0.9%) |  |
| Satisfaction Reported | Excellent | 36471 (59.8%) | 6678 (21.1%) | 1383 (8.9%) | P<0.001 | 6778 (16.4%) | 21479 (49.7%) | 1438 (5%) | P<0.001 |
|  | Very Good | 19062 (31.3%) | 13953 (44%) | 2943 (18.9%) |  | 18223 (44%) | 17164 (39.7%) | 4440 (15.4%) |  |
|  | Good | 3986 (6.5%) | 8680 (27.4%) | 5117 (32.8%) |  | 13276 (32%) | 3944 (9.1%) | 9419 (32.7%) |  |
|  | Fair | 428 (0.7%) | 1698 (5.4%) | 4189 (26.8%) |  | 2708 (6.5%) | 362 (0.8%) | 9492 (33%) |  |
|  | Poor | 78 (0.1%) | 178 (0.6%) | 1628 (10.4%) |  | 173 (0.4%) | 27 (0.1%) | 3673 (12.8%) |  |
|  | Missing | 925 (1.5%) | 530 (1.7%) | 343 (2.2%) |  | 294 (0.7%) | 221 (0.5%) | 315 (1.1%) |  |

The characteristics of the clusters can be seen in Table-3 and within *S1 Table-6*. Within the ‘chronic pain cluster’ of both the hip and knee cohort a greater proportion of the patient’s report living alone, have higher rate of comorbidity, lower overall OJS scores, reduced improvement in the OJS. Furthermore, these patients have a lower EQ5D score when compared to the other clusters with a mean EQ5D nearly half that of other clusters (Table-3). Yet, many patients within the ‘chronic pain cluster’ following both hip and knee arthroplasty continue to report a successful and satisfactory operation. Albeit, to a lesser degree than patients assigned to cluster 1 or 2.

Using an ROC analysis, an optimal threshold of 15.5 was identified to highlight the ‘chronic pain cluster’ after hip arthroplasty. Hierarchical clustering performed to assess internal consistency yielded a comparable threshold of 13.5. The ROC analysis of knee cohort identified an optimal threshold of 18.5 on the OKS-PS that below which would identify the ‘chronic pain’ cluster. Again, hierarchical clustering was performed as part of an assessment of internal consistency, which yielded a comparable threshold of 15.5 (*Supplementary Information and Code*).

### Comparison to Self-Reported Pain

#### Hip Arthroplasty

A threshold of 15.5 on the OHS-PS identified 47.8% and 69.8% of patients reporting severe or moderate postoperative pain, respectively (Table-4). In a binary classification where moderate and severe postoperative pain are grouped and mild, very mild and no pain are grouped our threshold of 15.5 on the OHS-PS has a classification accuracy of 85.7% [95% CI: 85.5 – 86.0%] with a 60.6% sensitivity [95% CI: 59.8-61.2%] and 91.0% specificity [95% CI: 90.8-91.1%] (Table-4).

**Table-4.**
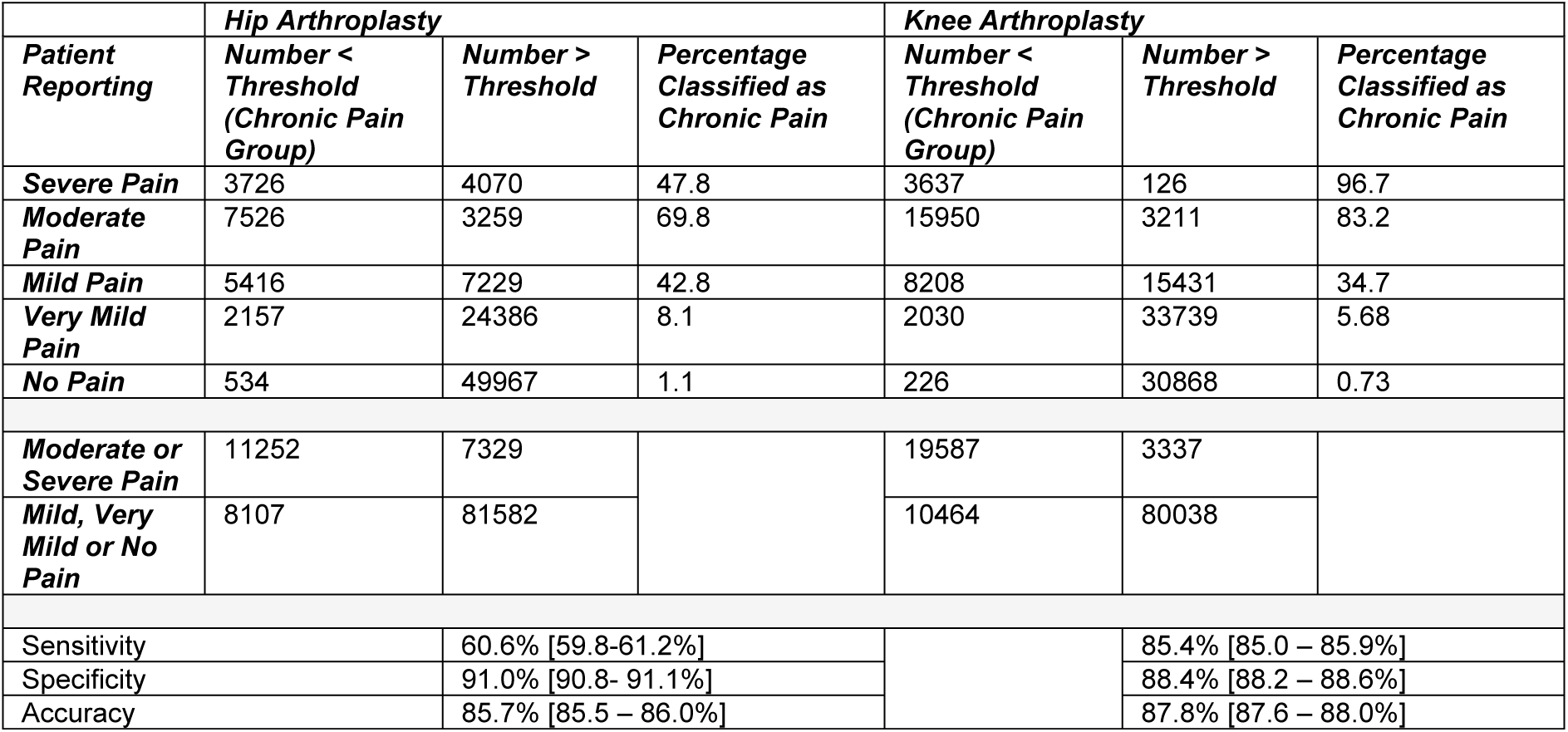
Summary of number of patients reporting varying severity of pain following either a hip or knee arthroplasty by threshold identified though cluster analysis. For hip arthroplasties a threshold on the OHS-PS of 15.5 was used, and for knee arthroplasties a threshold of 18.5 was used on the OKS-PS. The proportion of patients reporting pain and would be highlighted through use of our threshold is provided. *Below*: A binary classification of patient reported pain is provided where reporting moderate or severe pain are classified as chronic pain, accuracy, sensitivity and specificity are provided.

|  | *Hip Arthroplasty* | *Knee Arthroplasty* |
| --- | --- | --- |

| Patient Reporting | Number < Threshold<br>(Chronic Pain Group) | Number > Threshold | Percentage Classified as Chronic Pain | Number < Threshold<br>(Chronic Pain Group) | Number > Threshold | Percentage Classified as Chronic Pain |
| --- | --- | --- | --- | --- | --- | --- |
| Severe Pain | 3726 | 4070 | 47.8 | 3637 | 126 | 96.7 |
| Moderate Pain | 7526 | 3259 | 69.8 | 15950 | 3211 | 83.2 |
| Mild Pain | 5416 | 7229 | 42.8 | 8208 | 15431 | 34.7 |
| Very Mild Pain | 2157 | 24386 | 8.1 | 2030 | 33739 | 5.68 |
| No Pain | 534 | 49967 | 1.1 | 226 | 30868 | 0.73 |
| Moderate or Severe Pain | 11252 | 7329 |  | 19587 | 3337 |  |
| Mild, Very Mild or No Pain | 8107 | 81582 |  | 10464 | 80038 |  |
| Sensitivity |  | 60.6% [59.8-61.2%] |  |  | 85.4% [85.0 – 85.9%] |  |
| Specificity |  | 91.0% [90.8- 91.1%] |  |  | 88.4% [88.2 – 88.6%] |  |
| Accuracy |  | 85.7% [85.5 – 86.0%] |  |  | 87.8% [87.6 – 88.0%] |  |

#### Knee Arthroplasty

A threshold of 18.5 on the OKS-PS identified 96.7% and 83.2% of patients reporting severe or moderate postoperative pain, respectively (Table-4). This threshold has a classification accuracy of 87.8% [95% CI: 87.6 – 88.0%], with a sensitivity of 85.4% [95% CI: 85.0 – 85.9%] and specificity of 88.4% [95% CI: 88.2 – 88.6%] at discriminating between self-reported severe and moderate pain compared to mild, very mild or no pain (Table-4).

Fig-3 provides a visual representation of the questions making up the OJS-PS, divided using the thresholds identified following both hip and knee arthroplasty.

**Fig-3.**
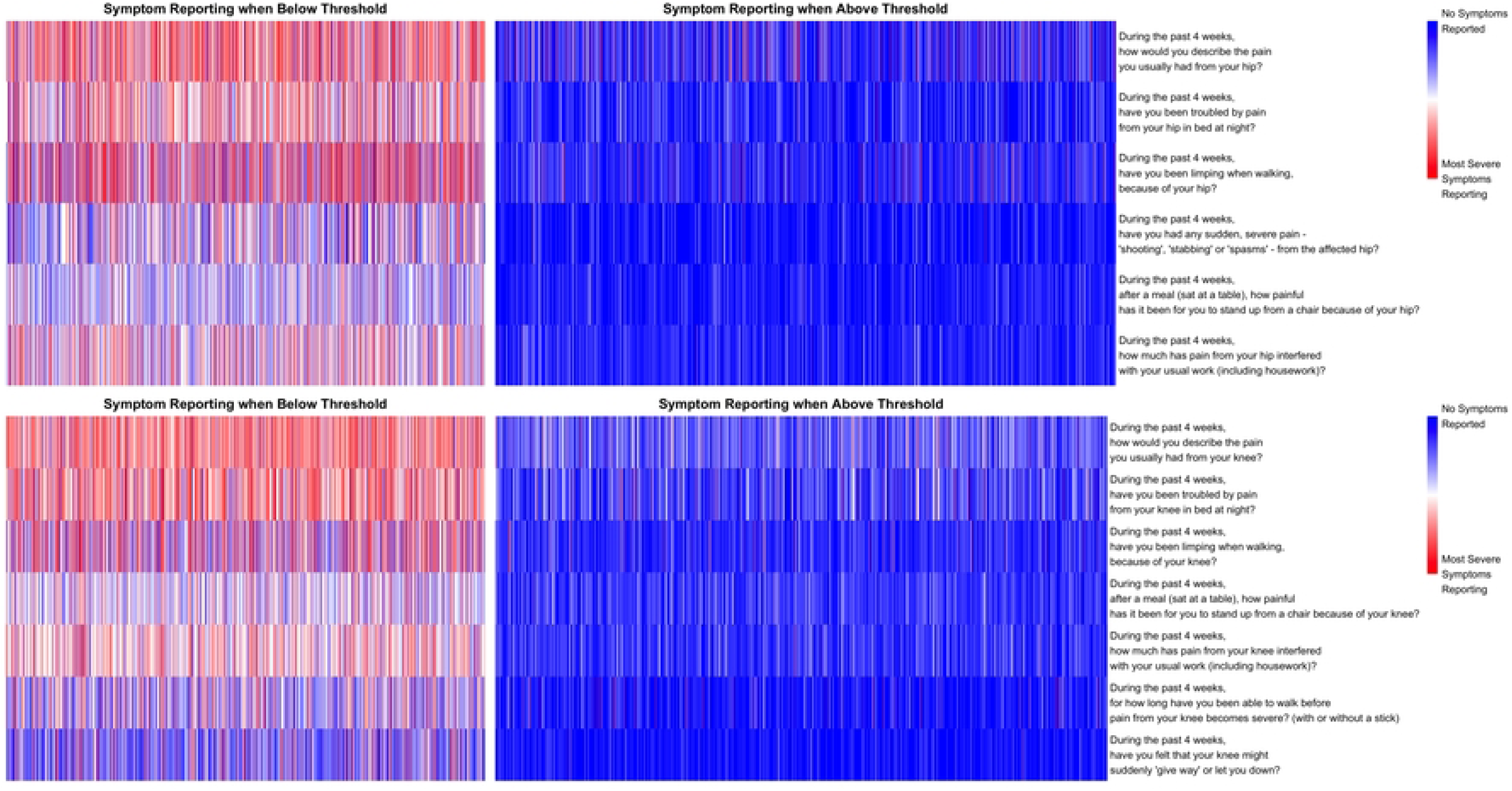
A heatmap summary of Oxford Joint Score when divided based on thresholds identified through cluster analysis. Colouring in red is indicative of poorer outcome reported by patients, whilst blue demonstrates better outcome reported by patients. Each row represents the question asked as part of the Oxford Joint score Pain Subscale. *Above*: answers following hip arthroplasty. *Below*: answers following knee arthroplasty.

## Discussion

Our robust and scalable *k*-medoids cluster analysis performed on ordinal non-ratio response data has identified a threshold on the OJS-PS that could aid screening of patients with a phenotype consistent with CPSP after hip or knee arthroplasty. To our knowledge this is the first time such a clustering algorithm has been used on clinical data with a sample size greater than 100,000. Patients with chronic pain may not present to healthcare services (15). As seen in Table-3, a proportion of patients within the ‘*chronic pain cluster’* report operative success and satisfaction – so may not present to healthcare services. From an epidemiological and public health perspectives utilising these thresholds at a population level could help screen for patients potentially with CPSP: consequently, improving diagnosis, treatment and outcome.

The proportion of patients with a phenotype consistent with CPSP was 14.4% following a hip arthroplasty and 25.3% following knee arthroplasty. These estimates are well within estimates of CPSP prevalence of between 7 and 23% following a hip arthroplasty and between 13 and 44% following a knee arthroplasty (10). Correspondingly, the proportion of patients within the ‘chronic pain cluster’ that live alone, have depression, or suffer with moderate anxiety is also higher (supplementary information S1 Table-6). Again, consistent with wider literature (10, 27). The ‘*chronic pain cluster’* patients have worse outcomes as measured by the EQ5D, OJS and satisfaction rating (Table-3). Being able to identify such patients and attempt to ‘*change course’* through assessment, diagnosis and treatment appears to be an important intervention, especially given the growing number of patients having both hip and knee arthroplasty performed (28, 29). Utilising our screening threshold, has the potential to identify such patients.

As previously noted, using a single response pain question to highlight chronic pain patients as this could be subject to recency bias, follow the peak-end rule, one-dimensional and does not reflect how chronic pain affects an individual’s life (30). Table-2 gives some indication of this, with a range of OJS-PS scores between different self-reported pain states: therefore implying difference in how the pain if effecting individuals lives and ability. There are some individuals whom report mild pain, yet continue to report poor outcomes in other pain domains, hence a low OJS-PS. Other individuals report severe pain, yet score a higher OJS-PS as other pain domains or function is not as impaired. Our clustering technique uses all responses as part of the OJS-PS, maintaining the multidimensionality of the pain assessment, with questions specifically noting symptoms from the weeks prior to the survey. In comparison to work by *Pinedo-Villanueva et al* (21), our threshold of 18.5 following a knee arthroplasty is within the range of thresholds they report. *Pinedo-Villanueva et al* report a threshold of 14 to identify chronic pain patients following a knee arthroplasty, whilst noting this threshold could be as high as 18.5. This is the first time such work has been undertaken following non-revisional hip arthroplasty. Furthermore, there is a difference in method, as we felt that pain reporting should not be handled as a numeric variable with ratio or interval properties. This is, to our knowledge, the first instance of this specific clustering technique being used in health research. Given the novelty, internal validity was added through a comparative assessment to patient self-reported pain. We demonstrated a high accuracy of classification between patients with an OJS-PS below threshold (potentially with a phenotype consistent with CPSP) and those that report moderate or severe postoperative pain at the same time (Table-4).

The implication of our sampling, batch-assignment and probabilistic assignment clustering technique with the Gower distance has widespread utility within health research given the growing use of large datasets with mixed data. So, whilst similar results have been obtained (21), the robust approach, scalability and implication for future healthcare research warrants the complexity of this approach.

### Limitations

Our analysis is based on survey data which can be prone to bias, either: due to sampling, non-response, self-selection or recall. Other issues include lack of distinction in surgical approach or anaesthetic technique, and patients can be included twice within the dataset.

This cross-sectional study is based on routinely collected data at 6 months postoperatively. Therefore, are unable to draw conclusions after 6 months. The trajectory of patients identified as phenotypically consistent with CPSP remains unknown. Conducting similar work on patients over time with repeated OJS-PS responses at 3, 6, 12, 18, 60 months would be preferential to determine a patient’s trajectory, or to explore how patients assigned to a ‘chronic pain cluster’ may change over time.

There needs to be caution around the adoption of thresholds into practice. Utility in clinical practice is limited, if a patient presents in pain use of this questionnaire with threshold is not going to change how a clinician treats and investigates. Limiting clinical practice based on a score and using as a diagnostic threshold would be unethical. However, there is utility as a population screening tool to identify patients who could be silently suffering.

### Future research

These thresholds are internally derived, therefore needs to be externally validated prior to any implementation within clinical practice. Ideally, through comparing the OJS-PS and our threshold with a more detailed assessment of chronic pain in a prospective manner. These thresholds, once validated, could be used to screen for potential CPSP patients at 6 months after arthroplasty or help in determining the clinical effectiveness of perioperative interventions aimed to reduce the prevalence of CPSP (31).

## Conclusion

Utilising our robust, scalable k-medoids cluster analysis has provided an estimate of the proportion of patients with a phenotype consistent with chronic postsurgical pain following a hip or knee arthroplasty. Cautiously utilising these thresholds could screen for patients who would not otherwise have presented and suffered. If externally validated this could aid in the evaluation of clinical interventions targeting CPSP after hip or knee arthroplasty.

## Author Contributions

LG conceptualisation; methodology; data curation; formal analysis; visualisation; writing – original draft; writing – review and editing.

BD formal analysis; supervision; writing – review and editing.

RK supervision; writing – review and editing.

JW supervision; writing – review and editing.

All authors reviewed and approved the final version of the manuscript.

## Data Availability

All data is freely available at https://digital.nhs.uk/data-and-information/data-tools-and-services/data-services/patient-reported-outcome-measures-proms code to collate datasets is supplied in supplementary material.

## References

1. Blom AW, Donovan RL, Beswick AD, Whitehouse MR, Kunutsor SK. Common elective orthopaedic procedures and their clinical effectiveness: umbrella review of level 1 evidence. BMJ. 2021;374:n1511.

2. Royal College of Surgeons of England. Surgery and the NHS in numbers. London: RCS England; 2014. Available from: https://www.rcseng.ac.uk/news-and-events/media-centre/media-background-briefings-and-statistics/surgery-and-the-nhs-in-numbers/.

3. Crawford RW, Murray DW. Total hip replacement: indications for surgery and risk factors for failure. Ann Rheum Dis. 1997;56(8):455–7.

4. National Joint Registry. 21st Annual Report 2024. London: National Joint Registry; 2024.

5. Heo SM, Harris I, Naylor J, Lewin AM. Complications to 6 months following total hip or knee arthroplasty: observations from an Australian clinical outcomes registry. BMC Musculoskelet Disord. 2020 Sep 10;21(1):602.

6. Salehi R, Alizadeh-Otaghvar H, Farhadi B, Najafi M, Torabi H, Hojjati H, et al. Prevalence of Surgical Site Infection After Hip Arthroplasty; a Systematic Review and Meta-Analysis. Arch Acad Emerg Med. 2024;12(1):e54.

7. Bagheri A, Sharifi Niknafs A, Farhadi B, Mazhari SA, Karimian P, Hekmati Pour N, et al. Incidence and Risk Factors of Surgical Site Infection After Knee Arthroplasty; a Systematic Review and Meta-Analysis. Arch Acad Emerg Med. 2025;13(1):e28.

8. Simon SJ, Patell R, Zwicker JI, Kazi DS, Hollenbeck BL. Venous Thromboembolism in Total Hip and Total Knee Arthroplasty. JAMA Netw Open. 2023;6(12):e2345883.

9. Schug SA, Lavand’homme P, Barke A, Korwisi B, Rief W, Treede RD, et al. The IASP classification of chronic pain for ICD-11: chronic postsurgical or posttraumatic pain. Pain. 2019;160(1):45–52.

10. Rosenberger DC, Pogatzki-Zahn EM. Chronic post-surgical pain - update on incidence, risk factors and preventive treatment options. BJA Educ. 2022;22(5):190–6.

11. Dueñas M, Ojeda B, Salazar A, Mico JA, Failde I. A review of chronic pain impact on patients, their social environment and the health care system. J Pain Res. 2016;9:457–67.

12. Varallo G, Giusti EM, Manna C, Castelnuovo G, Pizza F, Franceschini C, et al. Sleep disturbances and sleep disorders as risk factors for chronic postsurgical pain: A systematic review and meta-analysis. Sleep Med Rev. 2022;63:101630.

13. Turk DC, Fillingim RB, Ohrbach R, Patel KV. Assessment of Psychosocial and Functional Impact of Chronic Pain. J Pain. 2016;17(9 Suppl):T21–49.

14. Stark N, Kerr S, Stevens J. Prevalence and predictors of persistent post-surgical opioid use: a prospective observational cohort study. Anaesth Intensive Care. 2017;45(6):700–6.

15. Jinks C, Ong BN, Richardson J. A mixed methods study to investigate needs assessment for knee pain and disability: population and individual perspectives. BMC Musculoskelet Disord. 2007;8:59.

16. Redelmeier DA, Kahneman D. Patients’ memories of painful medical treatments: real-time and retrospective evaluations of two minimally invasive procedures. Pain. 1996;66(1):3–8.

17. Bendinger T, Plunkett N. Measurement in pain medicine. BJA Educ. 2016;16(9):310–5.

18. Harris KK, Price AJ, Beard DJ, Fitzpatrick R, Jenkinson C, Dawson J. Can pain and function be distinguished in the Oxford Hip Score in a meaningful way?: an exploratory and confirmatory factor analysis. Bone Joint Res. 2014;3(11):305–9.

19. NHS England. Patient Reported Outcome Measures (PROMs). 2026. Available from: https://digital.nhs.uk/data-and-information/data-tools-and-services/data-services/patient-reported-outcome-measures-proms.

20. Harris K, Dawson J, Doll H, Field RE, Murray DW, Fitzpatrick R, et al. Can pain and function be distinguished in the Oxford Knee Score in a meaningful way? An exploratory and confirmatory factor analysis. Qual Life Res. 2013;22(9):2561–8.

21. Pinedo-Villanueva R, Khalid S, Wylde V, Gooberman-Hill R, Soni A, Judge A. Identifying individuals with chronic pain after knee replacement: a population-cohort, cluster-analysis of Oxford knee scores in 128,145 patients from the English National Health Service. BMC Musculoskelet Disord. 2018;19(1):354.

22. Jakobsen JC, Gluud C, Wetterslev J, Winkel P. When and how should multiple imputation be used for handling missing data in randomised clinical trials - a practical guide with flowcharts. BMC Med Res Methodol. 2017;17(1):162.

23. Montelpare WJ, Read E, McComber T, Mahar A, Ritchie K. Applied Statistics in Healthcare Research: Theoretical concepts and computational methods using SAS applications and the Webulators. Charlottetown: Pressbooks; 2020.

24. Loftus TJ, Shickel B, Balch JA, Tighe PJ, Abbott KL, Fazzone B, et al. Phenotype clustering in health care: A narrative review for clinicians. Front Artif Intell. 2022;5:842306.

25. Steinley D. K-means clustering: a half-century synthesis. Br J Math Stat Psychol. 2006;59(Pt 1):1–34.

26. de Mathelin A, Cecchi NE, Deheeger F, Mougeot M, Vayatis N. OneBatchPAM: A Fast and Frugal K-Medoids Algorithm. Proc AAAI Conf Artif Intell. 2025;39(15):16172–80.

27. Fernández-de-Las-Peñas C, Florencio LL, de-la-Llave-Rincón AI, Ortega-Santiago R, Cigarán-Méndez M, Fuensalida-Novo S, et al. Prognostic Factors for Postoperative Chronic Pain after Knee or Hip Replacement in Patients with Knee or Hip Osteoarthritis: An Umbrella Review. J Clin Med. 2023;12(20):6624.

28. National Joint Registry. NJR StatsOnline: hospital procedure volumes. Hemel Hempstead: National Joint Registry; 2024. Available from: https://surgeonprofile.njrcentre.org.uk/Home/StatsIndex.

29. Matharu GS, Culliford DJ, Blom AW, Judge A. Projections for primary hip and knee replacement surgery up to the year 2060: an analysis based on data from The National Joint Registry for England, Wales, Northern Ireland and the Isle of Man. Ann R Coll Surg Engl. 2022;104(6):443–8.

30. Davidson MA, Tripp DA, Fabrigar LR, Davidson PR. Chronic pain assessment: a seven-factor model. Pain Res Manag. 2008;13(4):299–308.

31. Doleman B, Mathiesen O, Sutton AJ, Cooper NJ, Lund JN, Williams JP. Non-opioid analgesics for the prevention of chronic postsurgical pain: a systematic review and network meta-analysis. Br J Anaesth. 2023;130(6):719–28.

